# Introductions and evolutions of SARS-CoV-2 strains in Japan

**DOI:** 10.1101/2021.02.26.21252555

**Authors:** Reitaro Tokumasu, Dilhan Weeraratne, Jane Snowdon, Laxmi Parida, Michiharu Kudo, Takahiko Koyama

**Affiliations:** IBM Research- Tokyo, Tokyo, Japan; IBM Watson Health, Cambridge, MA 02142, USA; IBM TJ Watson Research Center, Yorktown Heights, NY 10598, USA

**Keywords:** SARS-COV-2, variant, COVID-19, Japan, quarantine, strains

## Abstract

COVID-19 caused by SARS-CoV-2 was first identified in Japan on January 15^th^, 2020, soon after the pandemic originated in Wuhan, China. Subsequently, Japan experienced three distinct waves of the outbreak in the span of a year and has been attributed to new exogenous strains and evolving existing strains. Japan engaged very early on in tracking different COVID-19 strains and have sequenced approximately 5% of all confirmed cases. While Japan has enforced stringent airport surveillance on cross-border travelers and returnees, some carriers appear to have advanced through the quarantine stations undetected. In this study 30493 genomes sampled in Japan were analyzed to understand the strains, heterogeneity and temporal evolution of different SARS-CoV-2 strains. We identified 12 discrete strains with a substantial number of cases with most strains possessing the spike (S) D614G and nucleocapsid (N) 203_204delinsKR mutations. 155 distinct strains have been introduced into Japan and 39 of them were introduced after strict quarantine policy was implemented. In particular, the B.1.1.7 strain, that emerged in the United Kingdom (UK) in September 2020, has been circulating in Japan since late 2020 after eluding cross-border quarantine stations. Similarly, the B.1.351 strain dubbed the South African variant, P.1 Brazilian strain and R.1 strain with the spike E484K mutation have been detected in Japan. At least 14 exogenous B.1.1.7 sub-strains have been independently introduced in Japan as of late March 2021, and these strains carry mutations that give selective advantage including N501Y, H69_V70del, and E484K that confer increased transmissibility, reduced efficacy to vaccines and possible increased virulence. Furthermore, various strains, which harbor multiple variants in the PCR primers and the probe developed by National Institute of Infectious Disease (NIID), are emerging. It is imperative that the quarantine policy be revised, cross-border surveillance reinforced, and new public health measures implemented to mitigate further transmission of this deadly disease and to identify strains that may engender resistance to vaccines.

## Introduction

SARS-CoV-2, the etiological agent of COVID-19 was first identified in Wuhan, China in late 2019 before rapid worldwide transmission in the first quarter of 2020. In just a year, the number of confirmed cases exceeded 148 million globally with 3.1 million deaths as of April 27^th^, 2021^1^ The actual number of infections are likely much higher by accounting for asymptomatic cases and mild disease that do not get tested and under reporting due to social stigma and discrimination associated with COVID-19 infections^2^. The novel coronavirus quickly spread to neighboring Japan, with a confirmed case of an individual on Jan 15, 2020 with travel history to Wuhan^3^. Soon after the identification of the first case, a major initial transmission catastrophe was averted when the government decreed the Diamond Princess cruise ship with infected patients to be anchored off the coast of Japan and mandated a 14-day quarantine for all passengers^4^. However, the number of confirmed infections has surpassed 572,000 with over 10 thousand reported fatalities in Japan^1^.

The COVID-19 pandemic has exerted an unprecedented stress on global health systems and has created a ripple effect touching every rubric of human life. Particularly the impact on healthcare (including mental health) and the underlying social, political, psychological and economic disruptions have had profound ramifications. In Japan, the dichotomy between the public health safety and societal and economic dynamics forced the postponement of the much-awaited summer Olympics and Paralympics games in Tokyo. Nonetheless, Japan implemented a stringent surveillance process at airports and seaports at the very early stages of the outbreak to monitor and quarantine travelers and repatriates with COVID-19 infection. In short, the rigorous surveillance process required a negative COVID-19 test prior to boarding, saliva antigen testing at the cross-border port-of-entry and, if positive, polymerase chain reaction (PCR) confirmation and sequencing at the National Institute of Infectious Diseases. While the surveillance process has been largely successful with 2392 patients detected and intercepted at quarantine stations by end of March 2021^5^, there appear to be some patients harboring exogenous strains with different haplotypes of the virus who were undetected at the port-of-entry. Japan has been vested and engaged from the beginning of the pandemic to monitor genomic changes in SARS-CoV-2 and has sequenced remarkable 30493 genomes which are approximately 5% of all confirmed cases. Notwithstanding the rigor of the public health measures, three discrete waves of the disease have been observed in Japan and it’s plausible that different viral strains may have contributed to each spike.

Mutations are inevitable as viruses evolve as a mechanism to cope with selective pressure and confer selective advantage. COVID-19 is the first pandemic to occur after inexpensive sequencing technologies became widely available. SARS-CoV-2 accumulates mutations at the rate of 1.0 x 10-3 (per site/genome/year), that corresponds to 2.5 mutations in a month^6^. However, as number of the infections increases, the number of variants increase proportionally. SARS-CoV-2 has accumulated and established multiple mutations within a year from the first published report of D614G in early April 2020^7^. New strains with spike N501Y mutation such as B.1.1.7 from England, B.1.351 from South Africa, P.1 from Brazil and R.1 have recently emerged^8^. Previously, N501Y has been functionally characterized and was reported to cause higher binding with ACE2^9^ and another study reported that B.1.1.7 carrying H69_V70del in addition to N501Y possess a higher infectivity rate of 75%^10^. Furthermore, B.1.351, P.1 and R.1 strains contain spike E484K and appear to confer reduced efficacy to the currently available vaccines^11,12^.

In this work we have evaluated the publicly available SARS-CoV-2 genomes in Japan to elucidate different viral strains that were exogenously introduced, to understand community transmission patterns and to delineate founder strains that further evolve within a community.

## Methods

Between 1 February 2020 and 5 April 2021, genomes were downloaded from the publicly available viral sequencing repositories, Global Initiative on Sharing All Influenza Data (GISAID)^13^, National Center for Biotechnology Information (NCBI), and NGDC Genome Warehouse, and the National Microbiology Data Center (NMDC); of the 939410 genomes downloaded, 30493 were from specimens collected in Japan. Low quality genomes with gaps and ambiguous bases over 50 base pairs in length (excluding at the start and the end of the genomes) were discarded. After completing quality control, variant analysis was performed on 638543 including 29679 Japanese genomes with the method described previously^6^. In brief, genomes were first aligned to the reference genome NC_045512 using The European Molecular Biology Open Software Suite (EMBOSS) needle^14^ with open gap penalty of 100 to filter out spurious frameshifts. Next, differences with the reference genome were extracted and annotated using gene definitions of SARS-CoV-2 as different variant types including missense, synonymous, non-coding and indels. All the obtained variants including global cases are stored in Supplemental Table S1. Subsequently, hierarchical clustering was performed on Japanese domestic cases to organize strains with similar haplotypes to construct a variant graph (Figure 1B).

**Figure 1.**
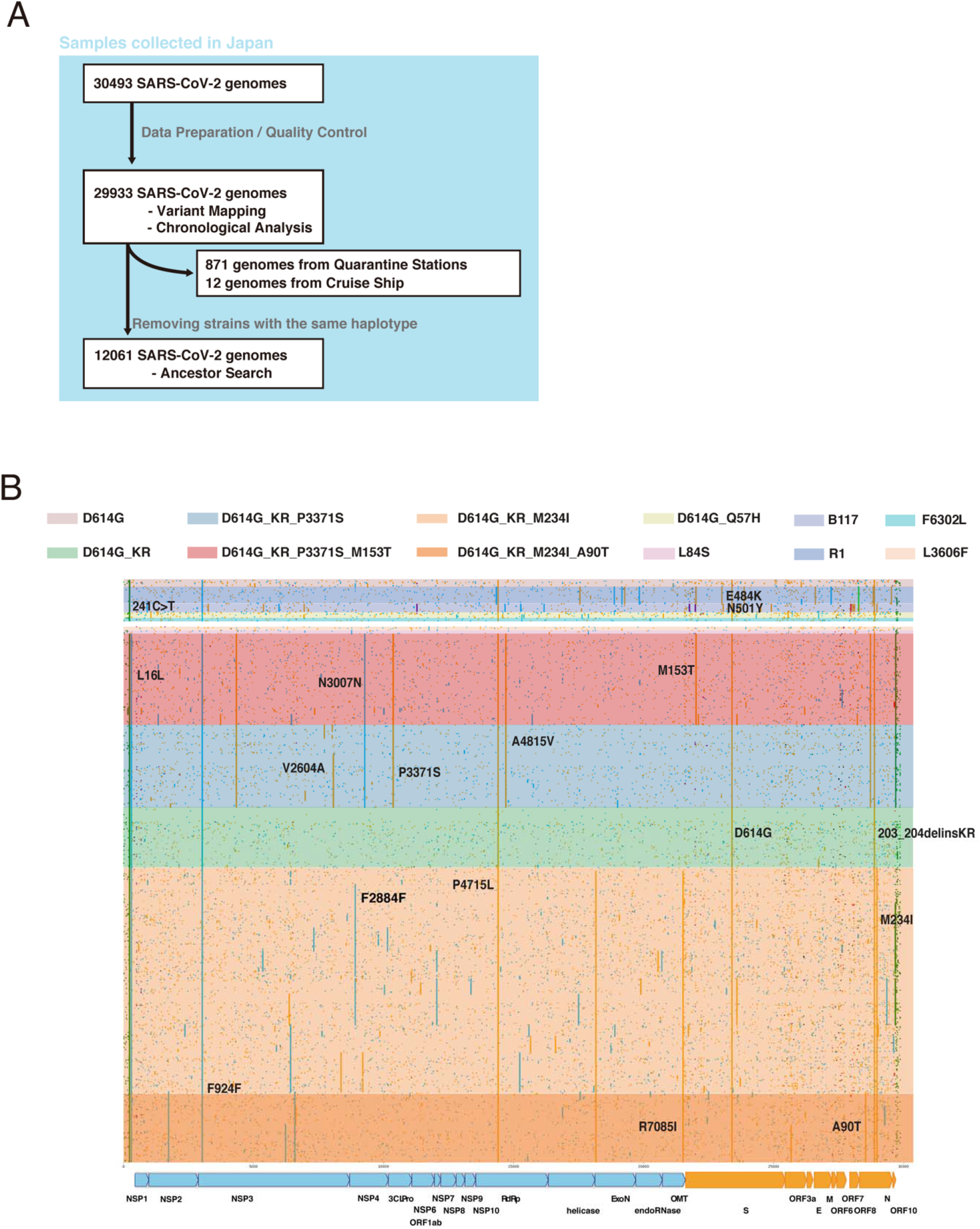
A Breakdown of Japanese SARS-CoV-2 genomes used in the study. B. Variant plot of SARS-CoV-2 strains in Japan. Missense variants are in orange, synonymous variants are in cyan, non-coding variants are in green and indels are in purple.

Haplotype defining major strains were extracted from the variant graph in Figure 1B. Numbers of confirmed cases and deaths in Japan obtained from Our World in Data^15^ were shown in Figure 2A. Monthly occurrences of each strain defined by the haplotype and its active period with evolutionary relationships are illustrated in Figure 2B.

**Figure 2.**
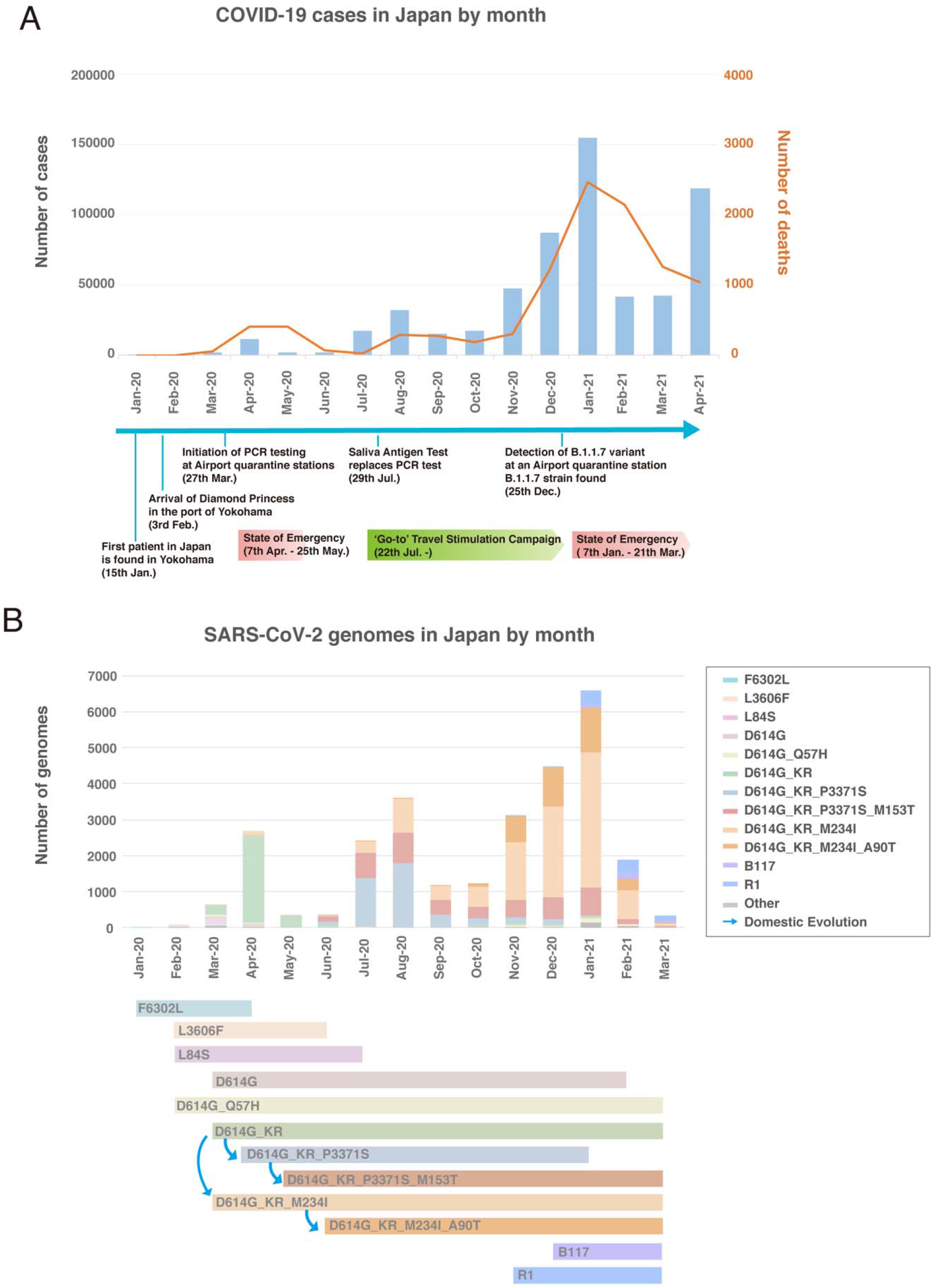
A. Statistics of cases and deaths in Japan from January 2020 to January 2021 with the event timeline. B. Monthly strains with classifications are plotted in the top and strains with the emergence and the last seen month are shown in the bottom. Evolutionary relationships are indicated with arrows.

A candidate parent of a particular strain was identified by the maximum variant approach using the haplotype of the query strain^16^. A parental strain should have a haplotype which is a subset of the query strain. Among ancestors obtained in the previous step, the closest ancestor is the one with maximal number of matches between haplotypes. The resulting parent and child relationships for all Japanese domestic cases are obtained in Supplemental Table S2.

Data on monthly COVID-19 positive cases at airport quarantine centers in Japan and number of passengers though immigration were obtained from a report released by the Japanese Government^5,17^ (Figure 4A). For each strain, we sought to identify a plausible parent including exogenous strains, which share more common variants than any domestic strains. Number of exogenous strains for each month is represented in Figure 4B. Among the exogenous strains, strains whose all probably ancestors were found after April 15th were considered to have advanced through quarantine stations undetected (Table 2).

**Table 1.**
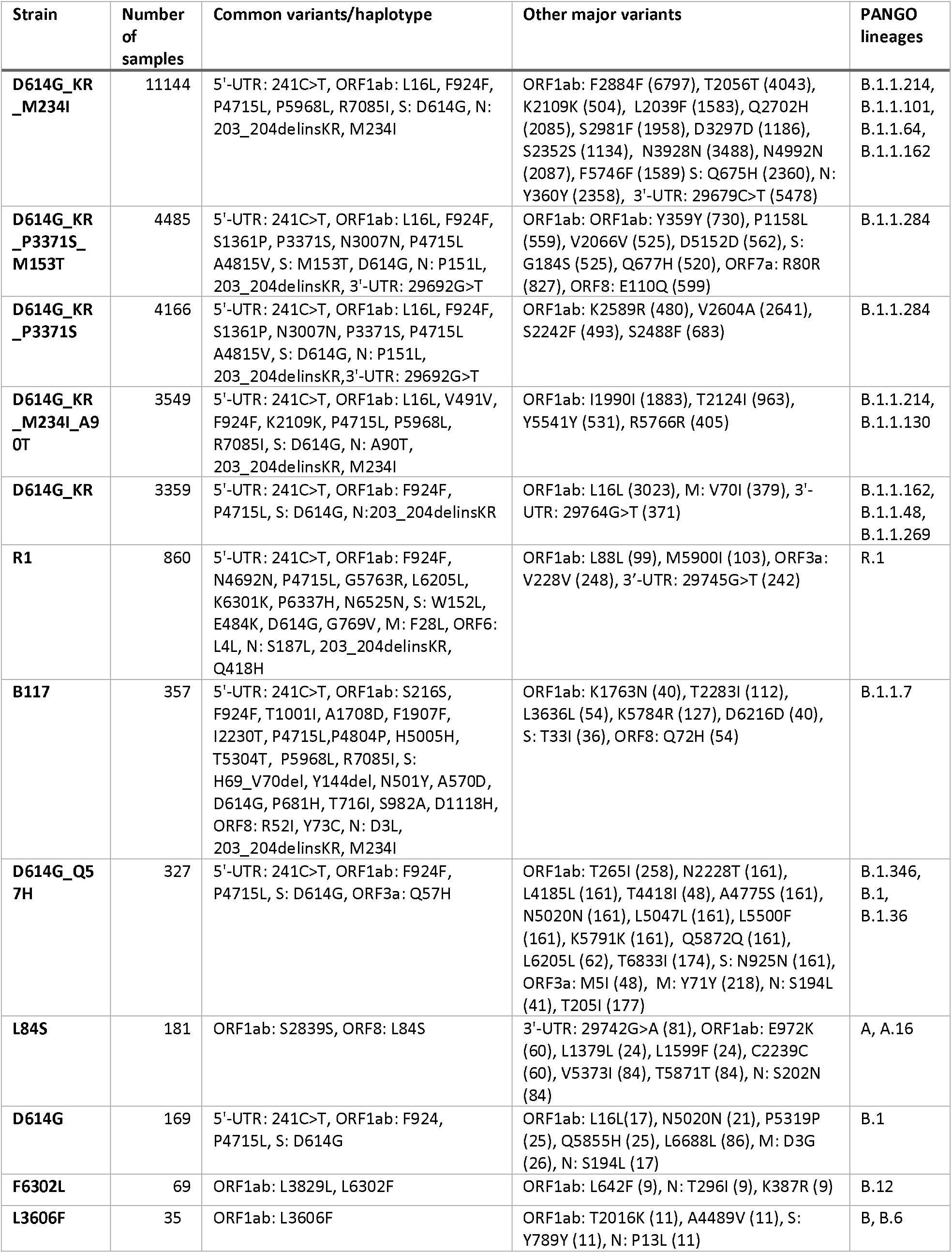
Major strains of SARS-CoV-2 circulating in Japan.

**Table 2.** List of strains which are likely introduced into Japan after quarantines were reinforced in March 2020. EPI_ISL_675164 does not meet our quality criteria with long undetermined bases.

| accession | Collection date | Earliest Ancestors in global samples | description |
| --- | --- | --- | --- |
| EPI_ISL_687718 | 2020-06 | EPI_ISL_553452 (United Kingdom, 2020-06-11) | D614G_KR (B.1.1.1) |
| EPI_ISL_686881 | 2020-07 | EPI_ISL_553921 (United Kingdom, 2020-06-09) | D614G_KR (B.1.1.1) |
| EPI_ISL_902575 | 2020-09 | EPI_ISL_734689 (Belgium, 2020-05-01) | D614G_Q57H (B.1.36) |
| EPI_ISL_898269 | 2020-10 | EPI_ISL_437723 (Saudi Arabia, 2020-04-16) | D614G_Q57H (B.1.36) |
| EPI_ISL_1431450 | 2020-10-10 | EPI_ISL_671943 (France, 2020-07-09) | D614G_Q57H (B.1.160) |
| EPI_ISL_895182 | 2020-11 | MW070060 (USA, 2020-08-21) | D614G_Q57H (B.1.346) |
| EPI_ISL_896964 | 2020-11 | MW035371 (USA, 2020-08-05) | D614G_Q57H (B.1.2) |
| EPI_ISL_915408 | 2020-11-10 | EPI_ISL_481126 (India, 2020-05-22) | D614G_KR (B.1.1.269) |
| EPI_ISL_1432983 | 2020-11-30 | EPI_ISL_678644 (United Kingdom, 2020-11-03) | D614G_KR (B.1.1.216) |
| EPI_ISL_893239 | 2020-12 | EPI_ISL_777283 (United Kingdom, 2020-12-17) | B.1.1.7 |
| EPI_ISL_892701 | 2020-12 | EPI_ISL_811496 (United Kingdom, 2021-01-02) | B.1.1.7 |
| EPI_ISL_892703 | 2020-12 | EPI_ISL_629440 (United Kingdom, 2020-10-22) | B.1.1.7 |
| EPI_ISL_1131416 | 2020-12-28 | MW523460 (USA, 2020-11-23) | D614G_KR (B.1.1.222) |
| EPI_ISL_1426529 | 2020-12-28 | EPI_ISL_605802 (India, 2020-05-01) | D614G_KR (B.1.1.216) |
| EPI_ISL_1429167 | 2020-12-29 | EPI_ISL_848297 (USA, 2020-07-21) | D614G_Q57H (B.1.2) |
| EPI_ISL_1072966 | 2021-01 | EPI_ISL_735562 (United Kingdom, 2020-12-13) | B.1.1.7 |
| EPI_ISL_1427595 | 2021-01-03 | EPI_ISL_629703 (United Kingdom, 2020-10-21) | B.1.1.7 |
| EPI_ISL_1128140 | 2021-01-07 | EPI_ISL_817111 (United Kingdom, 2020-12-18) | B.1.1.7 |
| EPI_ISL_1123466 | 2020-12-29 | EPI_ISL_675164 (United Kingdom, 2020-11-14)* | R.1 |
| EPI_ISL_1425748 | 2021-01-10 | EPI_ISL_675164 (United Kingdom, 2020-11-14)* | R.1 |
| EPI_ISL_1123723 | 2021-01-12 | EPI_ISL_1258396 (Canada, 2021-01-18) | D614G_KR (B.1.1.222) |
| EPI_ISL_1124365 | 2021-01-16 | EPI_ISL_1039729 (USA, 2021-01-19) | D614G_Q57H (B.1.2) |
| EPI_ISL_1425253 | 2021-02-08 | EPI_ISL_675164 (United Kingdom, 2020-11-14)* | R.1 |
| EPI_ISL_1425258 | 2021-02-08 | EPI_ISL_864200 (United Kingdom, 2021-01-12) | B.1.1.7 |
| EPI_ISL_1428639 | 2021-02-09 | EPI_ISL_1213196 (Brazil, 2021-01-26) | P.1 |
| EPI_ISL_1429597 | 2021-02-12 | MW583267 (USA, 2021-01-29) | D614G_Q57H (B.1.2) |
| EPI_ISL_1433297 | 2021-02-15 | EPI_ISL_1110039 (USA, 2021-01-29) | B.1.1.7 |
| EPI_ISL_1430660 | 2021-02-19 | EPI_ISL_1016856 (New Zealand, 2021-02-12) | B.1.1.7 |
| EPI_ISL_1430662 | 2021-02-19 | EPI_ISL_1016856 (New Zealand, 2021-02-12) | B.1.1.7 |
| EPI_ISL_1427683 | 2021-02-21 | EPI_ISL_977354 (Zambia, 2020-12-01) | D614G_Q57H (B.1.351) |
| EPI_ISL_1430736 | 2021-02-22 | EPI_ISL_1095590 (Turkey, 2021-02-08) | D614G_Q57H (B.1.351) |
| EPI_ISL_1425490 | 2021-02-24 | EPI_ISL_675164 (United Kingdom, 2020-11-14)* | R.1 |
| EPI_ISL_1425471 | 2021-02-25 | EPI_ISL_675164 (United Kingdom, 2020-11-14)* | R.1 |
| EPI_ISL_1430580 | 2021-02-26 | EPI_ISL_643356 (United Kingdom, 2020-10-29) | B.1.1.7 |
| EPI_ISL_1432765 | 2021-02-26 | EPI_ISL_768324 (Singapore, 2020-12-30) | D614G_Q57H<br>(B.1.466.2) |
| EPI_ISL_1431135 | 2021-02-27 | EPI_ISL_768324 (Singapore, 2020-12-30) | D614G_Q57H<br>(B.1.466.2) |
| EPI_ISL_1430659 | 2021-03-04 | EPI_ISL_1441714 (Germany, 2021-03-16) | B.1.1.7 |
| EPI_ISL_1430682 | 2021-03-05 | EPI_ISL_864619 (Switzerland, 2021-01-12) | B.1.1.7 |
| EPI_ISL_1433306 | 2021-03-06 | EPI_ISL_1441714 (Germany, 2021-03-16) | B.1.1.7 |

Strains which NIID_N qPCR probe fails to detect were identified by counting mutations in primers and probe sites. If any given strain has mutations involving multiple bases in the probe or in the primers, the strain was considered undetectable (Table 3).

**Table 3.** List of samples with multiple variants in primers and probe regions of NIID_N PCR tests

| Accession | country | collection date | mutations |
| --- | --- | --- | --- |
| EPI_ISL_1271520 | France | 2/26/2021 | 29224G>T, 29239G>A |
| EPI_ISL_1257834 | Indonesia | 1/26/2021 | 29225_29226delinsGA |
| EPI_ISL_1469240 | Indonesia | 2/18/2021 | 29225_29226delinsGA |
| EPI_ISL_1257855 | Indonesia | 2/5/2021 | 29224_29225delinsCA,<br>29229_29232delinsTATC |
| EPI_ISL_789044 | Sweden | 12/28/2020 | 29234_29235delinsTT |
| EPI_ISL_648206 | Sweden | 9/28/2020 | 29234_29235delinsTT |
| EPI_ISL_707699 | Tunisia | 11/23/2020 | 29234_29235delinsTT |
| EPI_ISL_707793 | Tunisia | 4/11/2020 | 29234_29235delinsTT |
| EPI_ISL_830792 | Switzerland | 2020 | 29234_29235delinsTT |
| EPI_ISL_1020124 | Spain | 1/29/2021 | 29250_29266del |
| EPI_ISL_1307717 | United States | 2/18/2021 | 29266_29301del |
| EPI_ISL_1089783 | Jordan | 1/29/2021 | 29257_29314del |

## Results

30493 genomes obtained between January 2020 and April 2021 were used for this analysis, accounting for about 5% of the confirmed cases in Japan (Figure 1A). We identified that frequently observed variants are shown in variant graph (Figure 1B). Spike protein D614G followed by ORF1ab P4715L (RdRp P323L), 5’-UTR 241C>T, nucleocapsid protein (N) 203_204delinsKR and ORF1ab L16L (313C>T) are the most common variants among strains in Japan, representing over 90% of all variants. L16L is unique to Japanese D614G_KR and its derivative strains while other three variants are common to all global D614G clade derived strains. Among the top non-synonymous variants, ORF1ab S1361P, P3371S, A4815V, S1261P, P5968L, R7085I, N P151L, M234I and spike M153T, are highly specific to Japanese strains. P3371S, which is located in 3CLPro, is reported to reduce the protease activity^18^.

By and large, strains circulating in Japan belong to one of the 12 strains as shown in Table 1; number of samples belong to the strain, defining haplotype, and other common variants are displayed with associated PANGO lineages^19^. The most frequently observed strain is D614G_KR_M234I, which harbors P5968L, R7085I, and N: M234I and most likely evolved from D614G_KR in Japan in March 2020. Figure 2A depicts number of positive cases and deaths during the period along with important events. As shown in Figure 2B, the Wuhan strain (wild type strain) and F6302L strain were introduced into Japan in January 2020 followed by L84S, and L3606F strains in February. In March, D614G related strains D614G, D614G_KR, D614G_Q57H were introduced into Japan. In April, a surge in D614G_KR occurred along with the emergence of D614G_KR_P3771S. In May, D614G_KR_P3771S_M153T emerged from the parental strain D614G_KR_P3771S and started gaining momentum during the summer. Although D614G_KR_P3371S_M153T was first observed in May, transmission route does not originate from a single founder(Figure 3). Therefore, the actual emergence appears to be at an earlier time. Significant decrease in numbers were observed in June marking the end of the first wave. In July, Japan experienced the second wave of the pandemic with three predominant strains detected: D614G_KR_P3771S, its offspring D614G_KR_P3771S_M153T, and D614G_KR_M234I. Although infection rates plateaued in October, Japan entered the third wave in November. While there is a paucity of available genomes in November, D614G_KR_M234I appears to be the dominant strain in the third wave. R1 and B117 strains were introduced in late 2020 and started gaining momentum in the fourth wave replacing existing strains.

**Figure 3.**
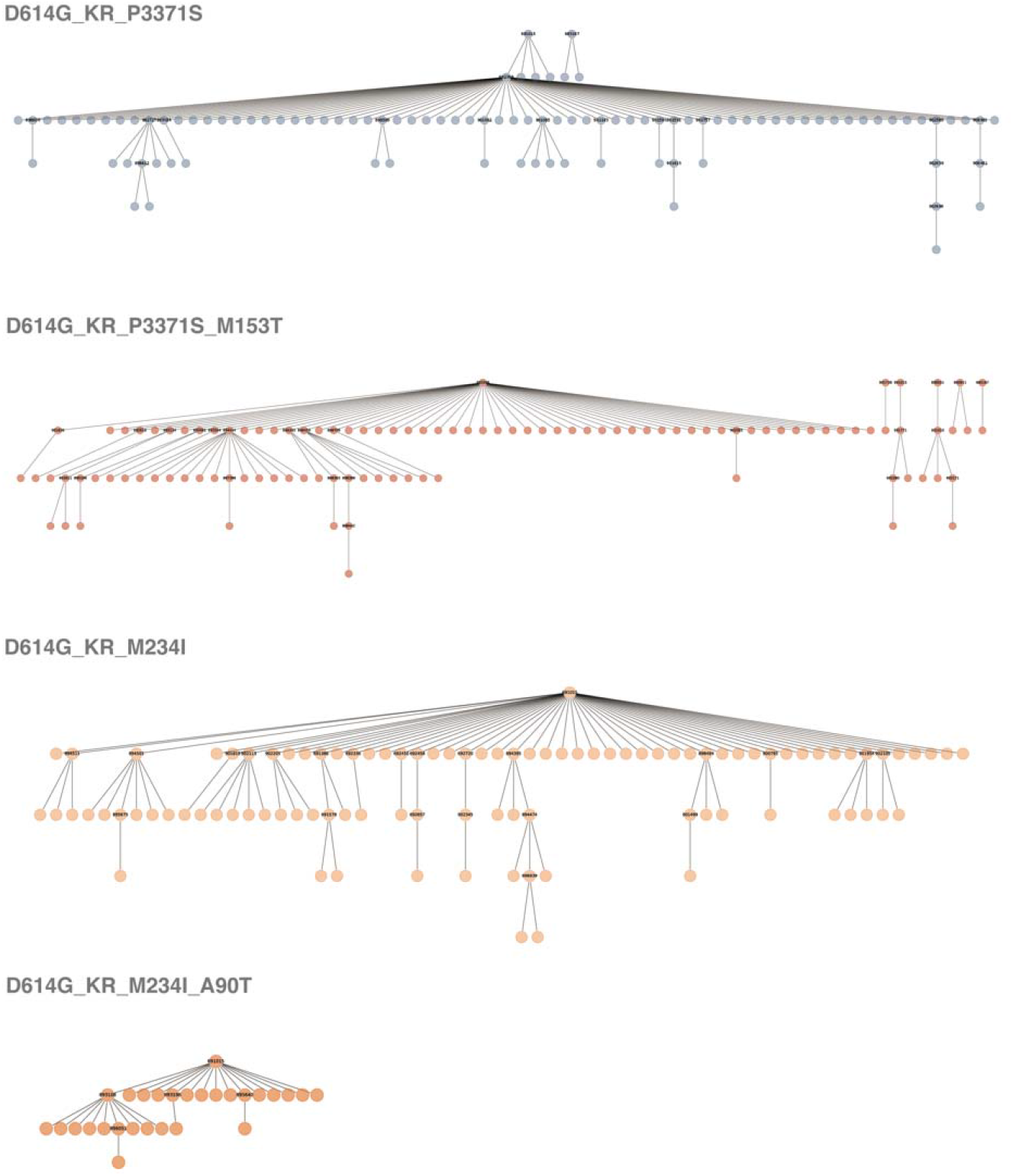
Transmission trees of Japan specific strains, D614G_KR_P3371S, D614G_KR_M234I, D614G_KR_P3371S_M153T, and D614G_KR_M234I_A90T are shown. For strains with more than 5 children are labeled with accession number and gained non-synonymous variants.

**Figure 4.**
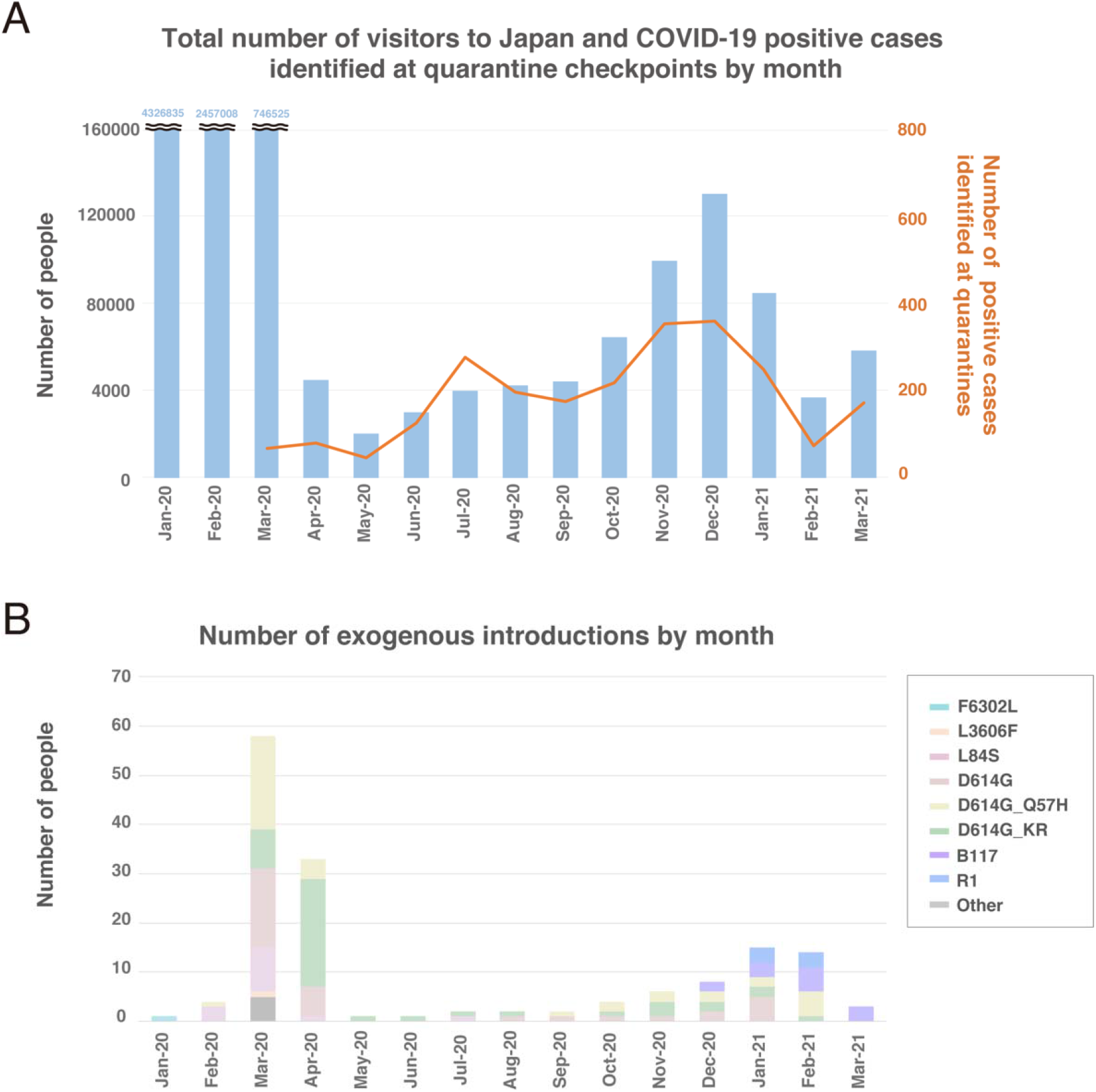
A. Number of inbound travelers are plotted with number of positive cases found at airport quarantine stations. B. Number of identified exogenous strains in each month are shown with strain classifications.

2392 positive cases were detected and intercepted at airport quarantine stations by the end of March 2021 since its inception in March 2020; of which 871 samples were sequenced(Figure 4A). In total, we have identified that 155 distinct strains have been introduced into Japan. However, the number of potentially undetected cases that evaded the surveillance scrutiny during the same time period was 39 after eliminating ambiguous cases as shown in Table 2. Most of the exogenous strains appears to have been introduced into Japan before strict airport surveillance policy was implemented as shown in Figure 4B. Among the 39 cases, 14 independent B.1.1.7 strains advanced through the quarantine checkpoints while 76 individuals carrying the strains were intercepted and quarantined. Besides spike N501Y variant, it is quite concerning that spike E484K mutation has been reported to reduce vaccine efficacy^12^. Furthermore, strains harboring spike E484K, namely, B.1.351, the South African strain, P.1, the Brazilian strain, and recently identified R.1 strain have been introduced into Japan. The R.1 strains were first observed in Niigata Prefecture in November (GISAID: EPI_ISL_895012); however, this strain has an extra mutation ORF1ab T265I compared with EPI_ISL_1123466, which is identified in late December as seen in Table 2. We were not able to identify the progenitor R.1 strain in any of the high-quality genomes in the cohort. However, we discovered a probable ancestor in a genome from United Kingdom (UK) (GISAID: EPI_ISL_675164) collected in November 14 which had long undetermined bases beyond the quality threshold of this study. To be exact, EPI_ISL_675164 has 4456C>T (ORF1ab A1397A) synonymous mutation, which Japanese R.1 strains are missing. UK has high volume of SARS CoV-2 sequencing, and no other R.1 strain was uncovered in November in UK. Furthermore, a sample from a Japanese quarantine station obtained from a traveler from Liberia in December (GISAID: EPI_ISL_736897) belongs to R.1. Combining these facts, it is likely that the R.1 strain may have originated elsewhere other than UK, where genomic surveillance is not frequently conducted.

Since qPCR tests have been performed to detect SARS-CoV-2, it is concerning that emerging strains are not detectable with qPCR tests. In Japan, NIID_N qPCR test kit was developed to detect SARS-CoV-2^20^. Although it is well designed and captures most of the strains reported today, there exists strains which harbor multiple mutations in primer and probe sites as shown in Table 3.

## Discussion

In this work we have identified multiple independently introduced discrete strains of SARS-CoV-2 in Japan which may have contributed to community spread. We also observed further evolution of exogenous founder strains after their introduction into Japan. It is quite intriguing that D614G_KR is dominant despite the fact that comparable numbers of D614G and D614G_Q57H strains were introduced into Japan. For unknown reasons, strains other than D614G_KR have not been able to establish and propagate in Japan^21^.

The airport quarantine stations have intercepted 2392 individuals carrying SARS-CoV-2 strains. For landlocked countries with contiguous borders, such quarantine strategy is not easy to implement. While cross-border surveillance is easier to implement in an island such as Japan, the study identified 39 cases, which had advanced through airport quarantine checkpoints and introduced to the Japanese population (Table 2). Since only 5% of confirmed cases have been sequenced, the actual number of individuals and SARS-CoV-2 strains passing undetected at quarantine stations is likely to be larger. Airport quarantine checkpoints have been utilizing antigen testing on saliva samples to improve lead times and throughput since July 27^th^ 2020. The number of positive cases detected at borders decreased in August in spite of the slight increase in number of people who entered Japan as seen in Figure 4A. It is more difficult to detect pre-symptomatic or asymptomatic patients with an antigen test on a saliva sample. It is concerning that the antigen test has low positive agreement rate of 55.2% with respect to qPCR resulting in high false negative rates for nasopharyngeal samples ^22^. In fact, four distinct B.1.1.7 strains have been identified and the number of people infected with B.1.1.7 has alarmingly augmented. Mandatory qPCR should be introduced to identify and intercept emerging strains such as B.1.1.7, B.1.351, P.1. and R.1. and any novel future strains. Additionally, global genomic data should be vigilantly monitored to identify new variants that potentially may be hard to discover with a traditional antigen or an existing qPCR test. As the virus accumulates more mutations, it is not possible to capture all the strains with a single probe test; therefore, it is advisable to utilize multiple probe qPCR.

Besides test accuracy at quarantine stations, compliance among travelers is another possibility of cases being undetected. Public health offices are urging cross-border travelers to be monitored for 14 days but there is no guarantee that they have followed the proper self-quarantine protocols. In fact, approximately 20% of cross border travelers have been lost for follow-up ^23^. Furthermore, some travelers enter Japan from exempt countries viewed as low risk; similarly, airline employees had been exempted from testing. During 14-day self-quarantine period, one can transmit to his/her family members or cohabitants, who can further transmit to people outside the household. Thus, it would be ideal to conduct interim and exit testing to further reduce infected passengers from circulating exogenous strains^24^.

There are several limitations of this study. While the genomes from airport quarantine checkpoints were released in a timely manner; the genomes from Japanese domestic samples lagged behind by months. Furthermore, more precise information on collection dates is needed; most of the Japanese samples only carry year and month but not the date. Moreover, specimen collection locations are not available on most instances. Therefore, it is challenging to evaluate spatial propagation of the virus and to assess the effect of a ‘go-to’ travel campaign to encourage business and leisure travel in an attempt to rescue the ailing travel industry. Therefore, while the study could not reveal unambiguous evidence, the campaign may have contributed to the resurgence of the virus across Japan and exacerbated the situation^25^.

It is imperative that the government mitigate the spread of new strains carrying the spike N501Y and E484K by reinforcing quarantine policies, further ramping up genome sequencing and large scale antigen testing for asymptomatic population^26^. In particular, wide spread of E484K variant strains may undermine vaccination efforts and discourage Japanese citizens from getting the vaccines. More aggressive genome sampling would facilitate understanding transmission dynamics of the virus including origins, routes, and rates, which is critical to its containment. Additional genomic analysis with clinical data as well as employing animal models will provide insights on infectivity, susceptibility, treatment resistance, vaccine evasion as well as the inferred pathogenicity and virulence^27,28^.

The COVID-19 pandemic is not over yet. Most citizens are not expected to be immunized by summer 2021, a time when Japan is hoping to host the postponed Olympic and Paralympics games. The onus is on the government of Japan to successfully executing these events while ensuring the public health safety of its citizens. The COVID-19 pandemic won’t be the last pandemic to affect mankind; therefore, reflections on the lessons learned through this ordeal should be used to ameliorate the effects of the next pandemic.

## Data Availability

All genome data used in the study is listed in supplemental table and available at GISAID and NCBI.
COVID-19 case and death number in japan is available at Our World in Data.

https://www.ncbi.nlm.nih.gov/

https://www.gisaid.org/

https://ourworldindata.org/

## Authors Contributions

T.K. conception of work. T.K. and R.T. acquisition and analysis of data. ALL interpretation of data. T.K. and R.T. drafted the work. D.W. and J.S. substantial revision. ALL reviewed the manuscript.

## Completing interests

The authors declare no competing interests.

## Acknowledgements

We gratefully acknowledge the authors, and the originating and submitting laboratories providing sequences from GISAID’s EpiFlu™ Database, NCBI and the National Microbiology Data Center (NMDC) is based. The list of genomes is provided in Supplemental Table S1.

## Supplemental Materials

Supplemental Table S1: Genomes Used in Analysis

Supplemental Table S2: Parent and child relationships among Japanese SARS-CoV-2 strains

